# Transition to multi-type mixing in d-dimensional spreading dynamics

**DOI:** 10.1101/2020.11.24.20238337

**Authors:** Alexei Vazquez

**Affiliations:** German Aerospace Center (DLR), Institute for the Protection of Terrestrial Infrastructures, Rathausallee 12, 53757 Sankt Augustin, Germany

## Abstract

The spreading dynamics of infectious diseases is determined by the interplay between geography and population mixing. There is homogeneous mixing at the local level and human mobility between distant populations. Here I model spatial location as a type and the population mixing by intra- and inter-type mixing patterns. Using the theory of multi-type branching process, I calculate the expected number of new infections as a function of time. In 1-dimension the analysis is reduced to the eigenvalue problem of a tridiagonal Teoplitz matrix. In *d*-dimensions I take advantage of the graph cartesian product to construct the eigenvalues and eigenvectors from the eigenvalue problem in 1-dimension. Using numerical simulations I uncover a transition from linear to multi-type mixing exponential growth with increasing the population size. Given that most countries are characterized by a network of cities with more than 100,000 habitants, I conclude that the multi-type mixing approximation should be the prevailing scenario.

## I. INTRODUCTION

Homogeneous mixing is a cornerstone in the mathematical analysis of infectious disease outbreaks [1, 2]. In a fully mixed population we define the basic reproductive number *R*_0_ = *β/γ*, where *β* is the average rate of disease transmission from infected to susceptible individuals and *γ* is the recovery rate from the disease. When *R*_0_ < 1 the epidemic outbreak dies out, but it grows exponentially in time when *R*_0_ > 1.

Human populations are not fully mixed, calling into question the value of *R*_0_. There are different mixing patterns according to age, immunization status and the adherence to government recommendations. Nevertheless, these heterogeneous mixing patterns can be treated under a generalized mixing approximation. If we stratified individuals into multiple types, then we can model the infectious disease spread as a multi-type spreading process [3–5]. In multi-type spreading processes *R*_0_ is replaced by the largest eigenvalue *ρ* of the mixing matrix. When *ρ* < 1 the infectious disease outbreak dies out, but it grows exponentially in time when *ρ* > 1.

Geographical heterogeneity seems to challenge the mixing hypothesis [6–8]. When simulated agents are constrained to a ring with nearest neighbours transmissions, the total number of infected cases grows linearly in time [6]. Sub-exponential infectious dynamics has been reported for agent based simulations in urban settings as well [7]. There are two caveats though. First, the observation of non-exponential dynamics is not sufficient evidence to rule out the mixing hypothesis. I have shown that a combination of a high reproductive number with a truncation of the disease transmission chain yields a power law growth in the number of new infections [9]. The pre-lockdown phase of the COVID-19 outbreak is consistent with this power low growth in the number of new infections [10–12]. Second, there is no prove that geography cannot be harnessed under some mixing approximation.

Here I demonstrate that geographical heterogeneity can be modelled as a type. Focusing on large cities, countries like Cuba look like a string of cities, with an effective 1-dimensional topology (Fig. 1a). Other countries, like Germany, are better represented by a two-dimensional mesh, with an effective 2-dimensional topology (Fig. 1b). At this level of description, we can model the network of cities as a multi-type network, where each city is represented by a type and the mixing pattern between cities accounts for the human mobility between them. In the following I introduce the multi-type mixing approximation for type networks with a d-dimensional topology, derive analytical results and test them with numerical simulations.

**FIG. 1.**
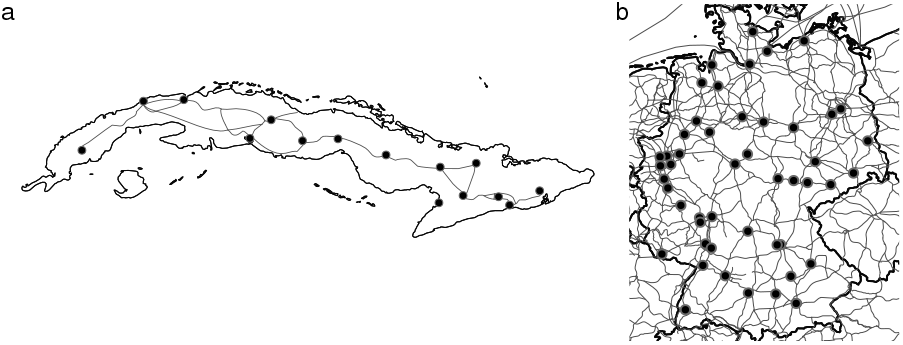
Geographical maps highlighting cities with more than 100,000 habitants and the road network connecting them. a) Cuba. b) Germany.

## II. MULTI-TYPE SIR MODEL

I investigate the susceptible, infected and removed (SIR) model on the multi-type network of cities. In the SIR model each individual is in one of three states: susceptible, infected and removed. Infected individuals transmit the disease to susceptible individuals, the latter becoming infected. Infected individuals recover or die from the disease, at which point they are removed from the disease transmission chain, the removed state.

### A. One-type branching process

When all individuals are of one-type we have a fully mixed population. We need to make a distinction though, between patient zero and any other case. In more detail, each individual contact other individuals at some rate *ζ*. Each contact results in disease transmission with probability *p*, resulting in the effective disease transmission rate *β* = *ζp*. Finally, the infected individuals are removed, due to recovery, isolation or death, at a rate *γ*. Patient zero is selected at random and its typical disease transmission rate is ⟨*β*⟩, where ⟨…⟩ denotes the average over the variability of *β* across individuals. Therefore patient zero has the average reproductive number

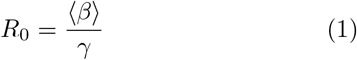

*R*_0_ is called the basic reproductive number.

For infected cases other than patient zero we take into account the disease transmission bias towards individuals with a higher contact rate. Any case other than patient zero is found with a probability proportional to its contact rate: *β/N*⟨*β*⟩, where *N* is the population size. Once infected, the individual found by contact will engage in new contacts at a rate *β*. Therefore, the average reproductive number for patients other than patient zero is

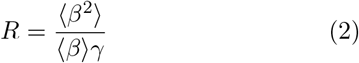

*R*_0_ gives the average number of infectious at the first generation, those generated by patient zero. *R*_0_*R* gives the average number of infections at the second generation and *R*_0_*R*^*k*−1^ gives the average number of infections at the *k* generation. The actual time when an infected case at generation *k* becomes infected equals the sum of *d* generation times

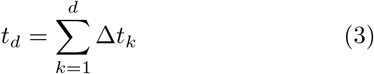

where Δ*t*_*k*_ are the generation intervals, the time interval from the infection of a primary case to infection of a secondary case in the transmission chain from patient zero. If the generation intervals have the probability density function *g*(*t*), then *t*_*d*_ has the probability density function *g*^*k*^(*t*), where the symbol ⋆ denotes convolution 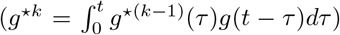. Putting the number of descendants and the timing together, we obtain the average number of new infected individuals at time *t*

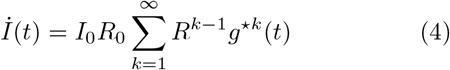

where *I*_0_ is the of number of patient zeros.

For the standard SIR model the distribution of generating intervals is the distribution of recovery times. Given that recovery takes place at a constant rate, the distribution of generation intervals is exponential

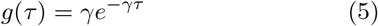

In this case, equation (4) becomes

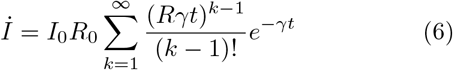

Noting that the series in (6) is the Taylor series expansion of the exponential, we obtain

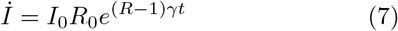

When *R* < 1 the disease dies out, but it grows exponentially when *R* > 1. *R* replaces the basic reproductive number *R*_0_ in the context of contact heterogeneity (⟨*β*^2^⟩ > ⟨*β*⟩2).

### B. Multi-type branching process

The multi-type formalism replaces the average reproductive number, a scalar, by a matrix of reproductive numbers. Let *n* be the number of types, representing cities or subpopulations. To patient zero we assign the reproductive number matrix *R*_0*n,n*_. The matrix element *R*_0*a,b*_ represents the average number of cases of type *b* generated by a patient zero of type *a*. To any other case we assign the reproductive number matrix *R*_*n,n*_. The matrix element *R*_*a,b*_ represents the average number of cases of type *b* generated by a case of type *a* that is not a patient zero. I have previously calculated the expected number of infected individuals of epidemic outbreaks in heterogeneous populations with a multi-type structure [4]. Although the calculation is more involved it yields an expression with the same structure as the one-type case

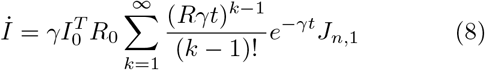

where *I*_0_ is a column vector representing the number of patient zeros by type and *J*_*n*,1_ is a column vector of ones. Or, making use of the exponential of a matrix,

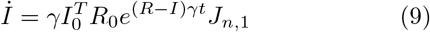

where *I* is the identity matrix.

### C. Diagonalizable *R*

When *R* is diagonalizable we obtain results that resemble the exponential dynamics of the single-type case.

Let *P* be the transformation matrix diagonalizing *R*,

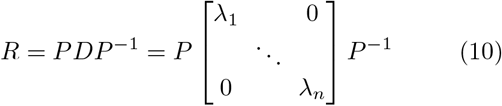

where *λ*_1_, …, *λ*_*n*_ are the eigenvalues of *R*. Note that some *λ*_*n*_ may be equal if some eigenvalues have multiplicity larger than 1.

I assume that *R*_0_ has the same form of *R*

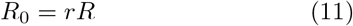

where the factor

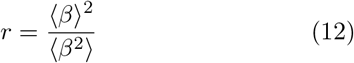

is the ratio between equations (1) and (2).

Finally, I will use the standard notation for functions of diagonal matrices

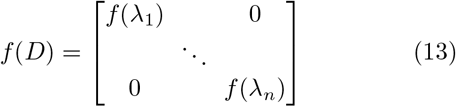

Using (10) and (11) we can rewrite (9) as

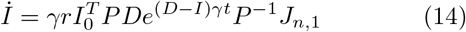

Whether the number of new cases grows or decays is determined by the largest eigenvalue

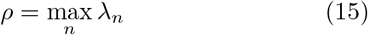

When *ρ* < 1 the disease dies out, but it grows exponentially when *ρ* > 1. *ρ* replaces the basic reproductive number in the context of multi-type mixing.

The predictive value of *ρ* can be extended beyond diagonalizable *R*. In most realistic scenarios there is a bi-directional path between every pair of cities in the network. In the language of graph theory, the network of cities is strongly connected. In such a case *R* is irreducible in addition to be non-negative. Using the Perron-Frobenious theorem for irreducible matrices [13], one can demonstrate that when *ρ* < 1 the disease dies out, but it grows exponentially when *ρ* > 1 [3, 4].

The diagonalization procedure is extended to any generation interval distribution. Re-starting from equation (4), without specifying the form of *g*(*t*), we obtain

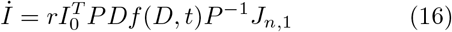

where

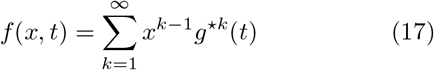

For the SIR model we have *g*(*t*) = *γe*^−*γt*^, *f* (*x, t*) = *γe*^(*x*−1)*γt*^ and we recover equation (14). I have calculated *f* (*x, t*) in terms of analytical functions for the gamma distributions of generation intervals [14]. Here I restrict the analysis to the SIR model. The extensions to other generation interval distributions is obtained after plugging in the specific function *f* (*x, t*).

## III. 1-DIMENSION

Now I analyze 1-dimensional topologies. Countries like Cuba have a 1-dimensional typology (Fig. 1a). In this case the reproductive number is given by

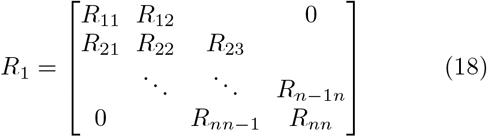

where *R*_*ii*_, *R*_*i*−1*i*_ and *R*_*ii*+1_ are the intra- and inter-city reproductive numbers. To obtain analytical results, I will work with homogeneous cities and left-right symmetric exchanges: *R*_*ii*_ = *a* and *R*_*i*−1*i*_ = *R*_*ii*+1_ = *b*. In this case *R* is a symmetric tridiagonal Toeplitz matrix.

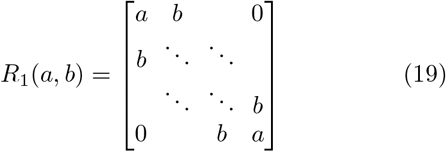

The eigenvalue problem of a tridiagonal Toeplitz matrix is solved exactly [15]. The eigenvalues are

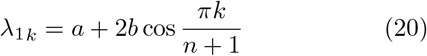

where *k* = 1, …, *n*. From the latter equation we obtain the largest eigenvalue

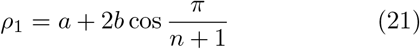

The intracity reproductive number *a* plus 2 times the intercity reproductive number *b*. The 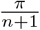 is a boundary correction. The type at the far left has no left neighbour and the type at the far right has no right neighbour.

Since 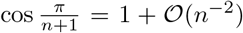. the boundary correction is irrelevant when *n* ≫ 1. The value of *ρ*_1_ can be used to determine whether the infectious disease dies out (*ρ*_1_ < 1) or grows (*ρ*_1_ > 1).

To account for all eigenvalues I go back to (14). The diagonalization matrix for the tridiagonal Toeplitz matrix (19) is given by [15]

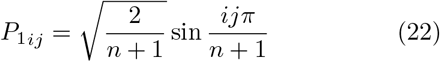

Since *R*_1_(*a, b*) in (19) is symmetric then *P* ^−1^ = *P*^*T*^. In this case (14) is simplified to

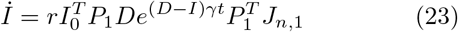

We can work directly with this equation. We can arrive to a more explicit expression as well. Substituting the form of *P*_1_ (22) and expanding the matrix sums we obtain

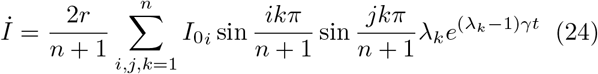

Using the trigonometric identity (AD 361.1 in Ref. [16])

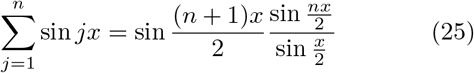

we can calculate the sum over the *j* index

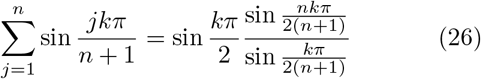

Substituting (26) into (24) and assuming a single patient zero at position *i*_0_ (*I*_0*i*_ = *δ*_*i,i*0_) we obtain

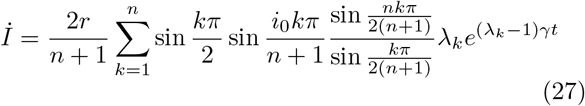

Figure 2 displays the analytical solution for a specific set of *a* and *b* values, together with the contribution of the largest eigenvalue alone. For *γt* > 10 the largest eigen-value is a very good approximation to the full analytical solution. Of note, I have double checked numerically that both (23) and (27) give the same result.

**FIG. 2.**
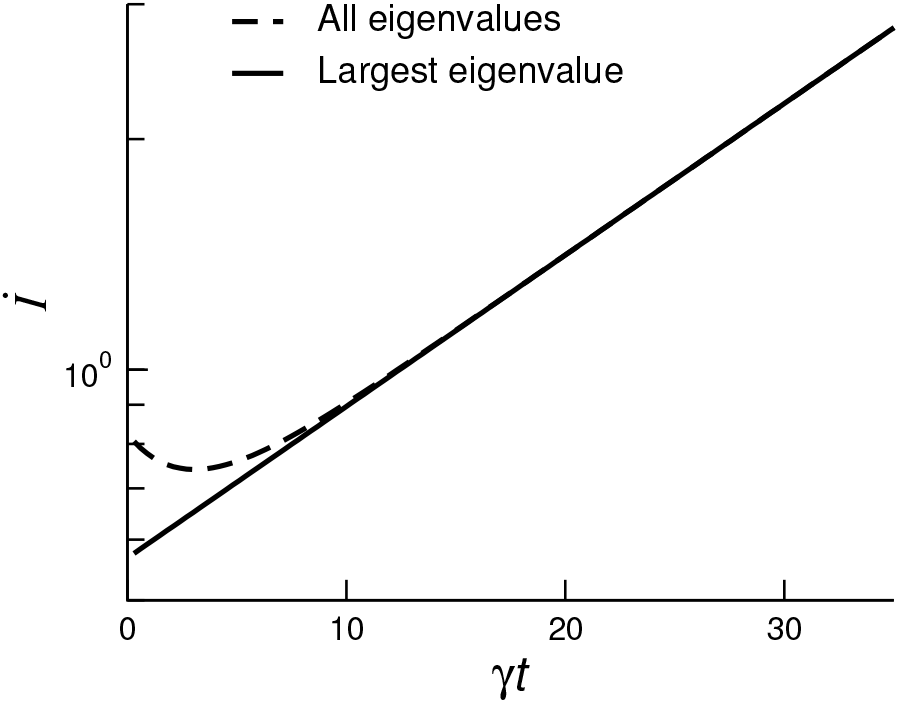
Multi-type dynamics in 1-dimension for the parameter set *r* = 1, *a* = 1.1*/*2, *b* = 1.1*/*4, and *n* = 6. The lines were generated with equation (24) using the contribution of all eigenvalues (dashed) or only the largest eigenvalue (solid).

## IV. 2-DIMENSIONS

In 2-dimensions I take advantage of the cartesian graph product and the 1-dimensional solution to calculate the eigenvalues. First, I introduce the definition of cartesian product of weighted graphs.

### A. Cartesian product of graphs

#### Cartesian product of graphs

Let *G* and *F* be a weighted graph with loops, adjacency-weighted matrices *A* and *B* and vertex count *n* and *m*, respectively. The cartesian product of *G* and *F*, denoted by *G*□*F*, is the graph with adjacency matrix

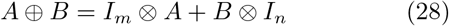

where *I*_*n*_ is the identity matrix of size *n*^2^, ⊕ denotes the Kronecker sum and ⊕ denotes the Kronecker product. The Kronecker product is defined as [17]

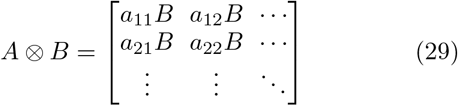

*Example:* An edge represents a complete simple graph with two nodes, denoted by *K*_2_. *K*_2_ has the adjacency matrix 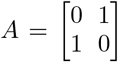 The cartesian product *K*_2_ □ *K*_2_ has the adjacency matrix *A* ⊕ *A* = *I*_2_ ⨂ *A* + *A* ⨂ *I*_2_

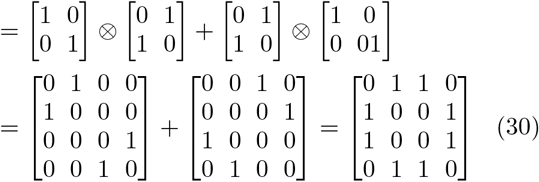

The latter is the adjacency matrix of a graph with vertices in a square. That is why the cartesian product is denoted by the symbol □.

#### Eigenvalues

The eigenvalues of the Kronecker sum equal the sum of the eigenvalues of the summed matrices [17]. If *α*_*k*_ and *β*_*l*_ are the eigenvalues of *A* and *B*, with associated eigenvectors *u*^(*k*)^ and *v*^(*l*)^ and transformation matrices *P* and *Q*, then

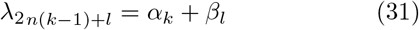

are the eigenvalues of *A* ⊕ *B*,

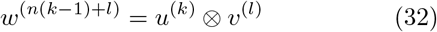

the associated eigenvectors and

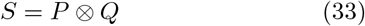

the transformation matrix diagonalizing *A* ⊕ *B*.

### B. 2-dimensional grid decomposition

We can decompose a 2-dimensional grid as the cartesian graph product between a 1-dimensional graph *G*_1_(*a, b*) with loops and another 1-dimensional graph *G*_1_(0, *b*) without loops (Fig. 3),

**FIG. 3.**
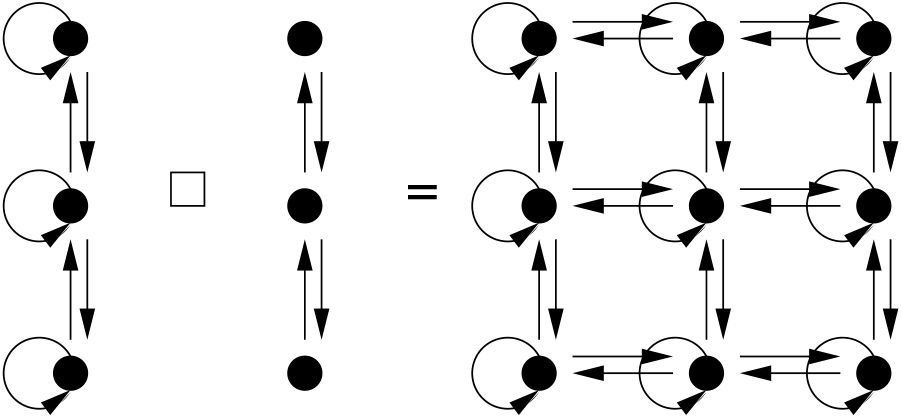
The cartesian product □ between a string with loops and a string without loops yields a 2-dimensional grid with loops.

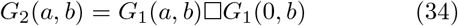

The adjacency matrix of *G*_2_(*a, b*) is

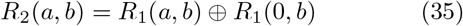

Substituting (20) into (31) we obtain the eigenvalues of *R*_2_(*a, b*),

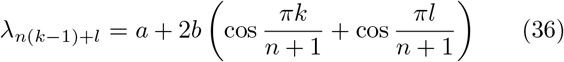

where *k* = 1, …, *n* and *l* = 1, …, *n*. From the latter equation we obtain the largest eigenvalue

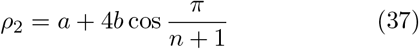

Noting that *R*_1_(*a, b*) and *R*_2_(0, *b*) are both diagonalized by *P*_1_, we substitute *P* = *Q* = *P*_1_ in (33) to obtain the transformation matrix in 2-dimensions

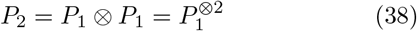

Substituting (38) into (14) and bearing in mind that 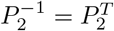 we obtain

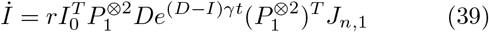

This equation is the multi-type mixing approximation for the average number of new infections in a 2-dimensional topology.

## V. *d*-DIMENSIONS

We can iterate the cartesian graph product to generate *d-*dimensional grids

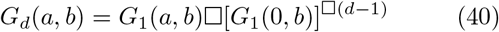

The average number of new infections in these *d*-dimensional grids is given by

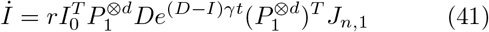

with eigenvalues

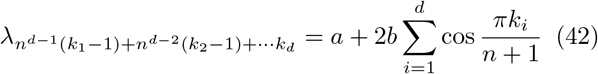

where *k*_*i*_ = 1, …, *n* and *i* = 1, …, *d*, and largest eigen-value

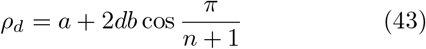

Applications for the 3-dimensional case include the spreading of prion protein aggregates in the human brain.

## VI. NUMERICAL SIMULATIONS

The basic reproductive number and its multi-type extension are defined in a contex where the number of infected individuals is much smaller than the population size. In a finite population the number of infected individuals may reach a significant percent of the population size. In that case the starting equation (4) is no longer valid. To investigate the validity of the analytical results and the population size effects, I will perform numerical simulations of the SIR model in 2-dimensional grids.

The cities are assumed of equal population size *H*. The number of susceptible individuals at city (*i, j*) is stored in the variable *S*_*ij*_. A list *L* is created with elements storing the coordinates of all infected individuals. The size of *L* is the number of individuals in the infected state, *I* =| *L* |. *γ* = 1 and time is measured in units of 1*/γ*.

*Initial conditions:* At *t* = 0 all individuals are in the susceptible state except from one individual at city (1, 1). *L* is initiated with the coordinates (1, 1) and *S*_*ij*_ = *H* for all (*i, j*) except for *S*_1,1_ = *H* − 1.

*Dynamics:* The Gillespie algorithm is used to update the system. At a total rate *µ* = (*a* + 4*b* + *γ*)*I* a new event happens. The time interval Δ*t* to this new even is extracted from the exponential probability density function *µe*^*µ*Δ*t*^. A coordinate is selected at random from *L* and stored in (*i, j*). With probability *γ/µ* the selected coordinate is removed from *L*. Otherwise, the disease transmission rule is applied as specified below. The time is updated *t*→ *t* + Δ*t*.

*Disease transmission:* With probability *a/*(*a* + 4*b*) we set (*i′, j′*) = (*i, j*), otherwise we set (*i′, j′*) equal to one of the 4 neighbours (*i* +1, *j*), (*i, j* +1), (*i, j* −1) and (*i*−1, *j*) with equal probability. With probability *S*_*i′*_, _*j′*_ */H* a susceptible individual in (*i′, j′*) gets infected, the coordinates (*i′, j ′*) are added to *L* and the updates *S*_*i′*_, _*j′*_ → *S*_*i′*_, _*j′*_ − 1 and *I* → *I* + 1 are performed. The empty boundary condition *S*_*i*,−1_ = *S*_*n*+1,*j*_ = *S*_*i,n*+1_ = *S*_−1,*j*_ = 0 are used.

*Statistics:* The number of new infections is recorded in time bins of size 1 and the average is calculated over multiple realizations.

### A. Subpopulation size effects

First I illustrate the transition to the multi-type branching approximation with increasing the cities population size. To this end I use the parameter set *r* = 1, *a* = 2 and *b* = 1*/*4. For *H* = 100 there is an evident linear increase of *İ* as a function of time (Fig. 4, circles). Furthermore, the numerical results are quite far from the multi-type mixing approximation (Fig. 4, line). For *H* = 1, 000 the numerical solution gets closer to the multi-type mixing approximation but it follows a linear growth (Fig. 4, squares). Yet, the linear range keeps reducing for *H* = 10, 000 and it is almost absent for *H* = 100, 000.

**FIG. 4.**
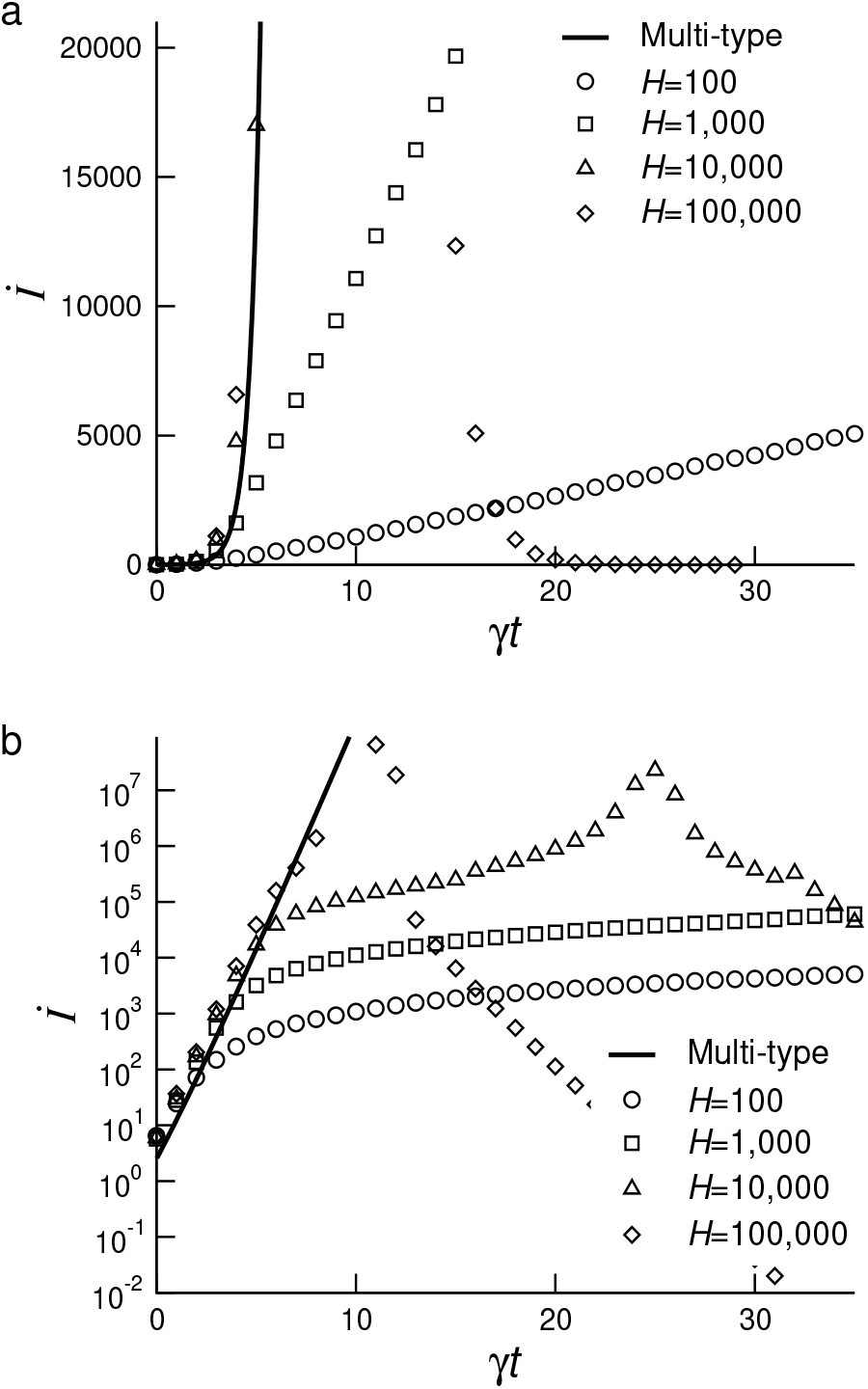
Average number of new infections for the SIR model in 2-dimensional grids with 100*×*100 cities, each with *H* habitants. The model parameters are *r* = 1, *a* = 2, *b* = 1*/*4. The average was calculated from 100 realizations. a) Y-axis in linear scale. b) Y-axis in log scale

As *H* increases we observe an increase in the region characterized by an exponential growth. Within the exponential growth regime the numerical solution coincides with the multi-type mixing approximation (Fig. 4, line).

### B. Lattice size effects

Second I report a lattice size effect. Based on equation (37), for *a*+4*b* ≳ 1 we can find scenarios where the reproductive number is smaller than 1 for *L* < *L*_*c*_ and larger than 1 otherwise. *L*_*c*_ is obtained by solving equation (37) for *L* with *ρ* = 1.

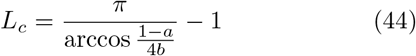

For example, for *a* = 1.1*/*2 and *b* = 1.1*/*4 the multi-type calculation predicts a transition from decay for *L* < 4.13 to growth for *L* > 4.13. Figure 5 reports the average number of new infections as a function of time for different values of cities size *H* = 100, 1,000, 10,000 and 100,000 and grid linear size *L* = 3, 4, 5 and 6. For *H* = 10, 000 and 100,000 there is confirmation of the multi-type mixing prediction. *İ* decays for *L* = 3, 4 (*L* < 4.13) while *İ* growths for *L* = 5, 6 (*L* > 4.13).

**FIG. 5.**
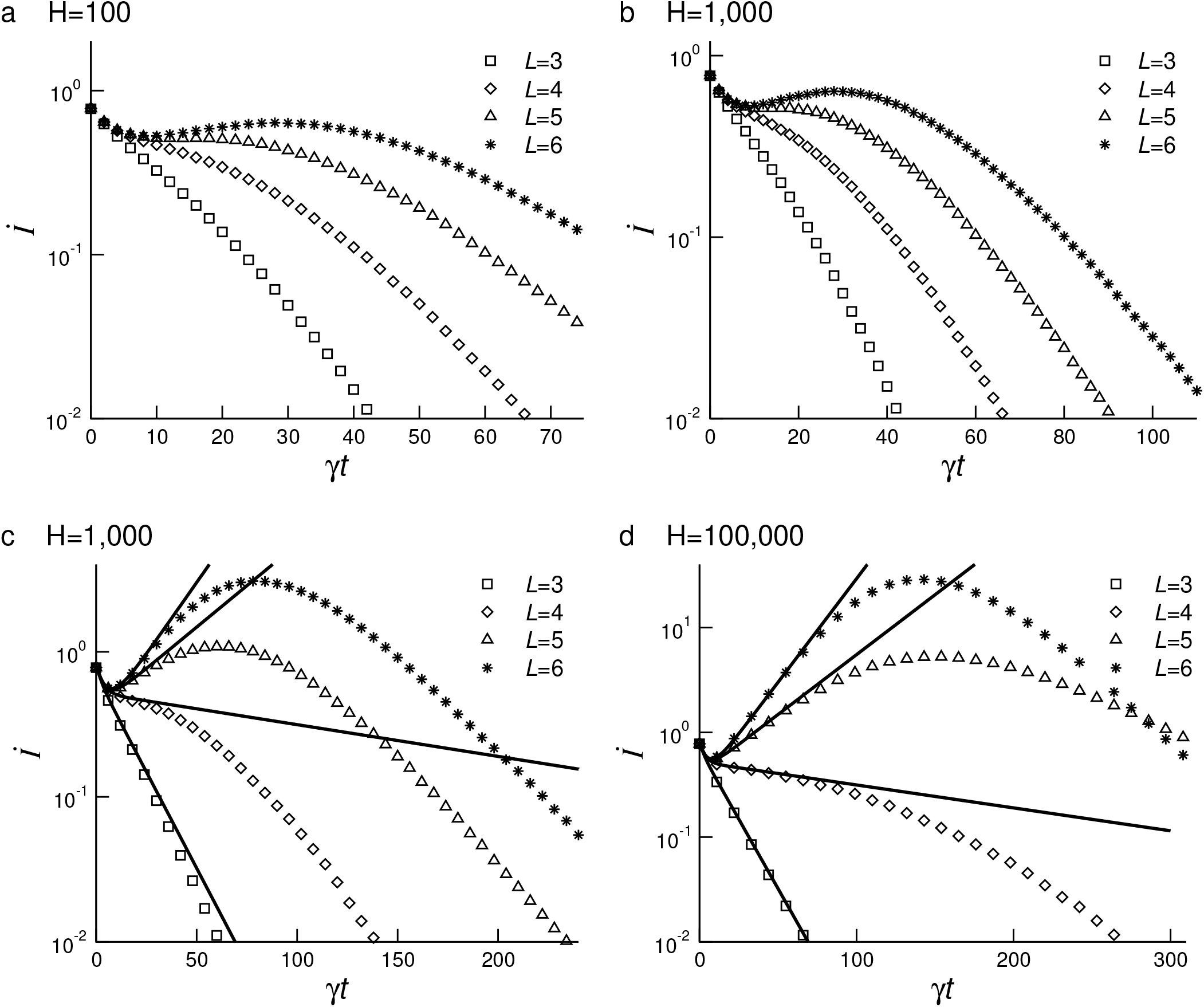
Average number of new infections for the SIR model in 2-dimensional grids with *n × n* cities, each with *H* habitants. The model parameters are *r* = 1, *a* = 1.1*/*2, *b* = 1.1*/*8 and *n* = *L*. The average was calculated from 1,000,000 realizations.

Here again there are city/subpopulation size effects. We do not observe the expected *İ* growth for *L* = 5 when *H* = 100 or 10,00. This example teach us that some diseases may not generate outbreaks when the network of cities is small, but as they city network grows they generate outbreaks.

For the largest city sizes *H* = 10, 000 and 100,000, I have plotted the analytical solution calculated from equation (41). At the early times the points from the numerical simulations fall in the multi-type mixing line. For longer times there is a deviation from the analytical results due to the finite population size.

## VII. CONCLUSIONS

I have demonstrated the use of the multi-type network approach and the cartesian product of graphs to calculate the epidemic threshold of spreading dynamics in *d*-dimensional grids. These calculations can be deployed to estimate the epidemic threshold at the country level aggregating cities/regions of the oder of 100,000 habitants or larger. The largest eigenvalue of the reproductive number matrix will provide a good estimate of the epidemic threshold. The analytical formula for the average number of new infections will provide a good approximation to the initial dynamics of the outbreak.

The numerical simulations in 2-dimensional grids uncovered a gradual transition from linear to exponential growth with increasing the cities/subpopulation size. The same is true for epidemic spreading in fully mixed populations. Depending on the magnitude of *R* and the implementation of lockdowns we can expect both exponential or power law growth [9]. Today countries are characterized by a network of cities with more than 100,000 habitants. In this context the numerical simulations reported here indicate an exponential growth of the number of new infections in time. Based on this observation I find unlikely that geography is the dominant factor behind the observation of power law growth. In contrast, the power law growth reported for COVID-19 is consistent with the time scales of the lockdown implementations [12]. However, I cannot exclude that there is a combination of both the lockdown truncation of the disease transmission chain and geographical location.

## Data Availability

All data have been reported in the manuscript.

